# Epigenetic scores of blood-based proteins as biomarkers of general cognitive function and brain health

**DOI:** 10.1101/2023.11.07.23298150

**Authors:** Hannah M. Smith, Joanna E. Moodie, Karla Monterrubio-Gómez, Danni A. Gadd, Robert F. Hillary, Aleksandra D. Chybowska, Daniel L. McCartney, Archie Campbell, Paul Redmond, Danielle Page, Adele Taylor, Janie Corley, Sarah E. Harris, Maria Valdés Hernández, Susana Muñoz Maniega, Mark E. Bastin, Joanna M. Wardlaw, Ian J. Deary, James P. Boardman, Donncha S. Mullin, Tom C. Russ, Simon R. Cox, Riccardo E. Marioni

## Abstract

**Background:** Blood-based biomarkers of brain health could provide a cost-effective contribution to detecting individuals at risk of dementia. Epigenetic scores (EpiScores) for blood protein levels have previously associated with several disease outcomes and measures of brain health, however this has typically been limited to single EpiScore analyse.

**Results:** Using 84 protein EpiScores as candidate biomarkers, associations with general cognitive function (both cross-sectionally and longitudinally) were tested in three independent cohorts: Generation Scotland (GS), and the Lothian Birth Cohorts of 1921 and 1936 (LBC1921 and LBC1936, respectively). A meta-analysis of general cognitive functioning results in all three cohorts identified 18 EpiScore associations (absolute meta-analytic standardised estimates ranged from 0.03 to 0.14, median of 0.04, FDR P<0.05). Several associations were also observed between EpiScores and global brain volumetric measures in the LBC1936. An EpiScore for the S100A9 protein (a known Alzheimer disease biomarker) was associated with general cognitive functioning (meta-analytic standardised beta: -0.06, P = 1.3 x 10^-9^), and with time-to-dementia in GS (Hazard ratio: 1.24, 95% confidence interval 1.08 – 1.44, P = 0.003), but not in LBC1936 (Hazard ratio: 1.11, P = 0.32).

**Conclusions:** EpiScores might make a contribution to the risk profile of poor general cognitive function and global brain health, and risk of dementia, however these scores require replication in further studies.

## Introduction

A projected 152 million people worldwide will have from dementia by 2050 (1). Dementia is characterised by cognitive decline with consequent serious limitations on performance of everyday activities, independence and quality of life in older age, even in the absence of dementia (2-4). Stable, consistent biological markers (biomarkers) of these outcomes might facilitate early detection, opening up a window for possible intervention (5). Biomarkers can also be used for monitoring progression, understanding the molecular mechanism of a phenotype, and identification of candidate drug targets. Proteins are commonly used as biomarkers, as changes in levels can be indicative of disease status or risk (6). Discovery of blood-based biomarkers is desirable as blood is easily accessible, can be taken at routine appointments and is cost-effective.

The term epigenetics refers to chemical modifications to DNA that do not affect the underlying sequence. The dynamic nature of these modifications can affect gene expression levels, therefore in turn affecting protein expression levels (7, 8). DNA methylation (DNAm) is the most commonly studied epigenetic modification, and is typically characterised by the addition of a methyl group to the cytosine base in a cytosine-guanine motif (CpG). Epigenetic scores (EpiScores) for proteins are typically derived from a linear weighted sum of DNAm sites that, in combination, are predictive of protein levels. A recent study directly compared measured CRP and EpiScore CRP levels, showing higher test-retest reliability for the EpiScore (9). For inflammatory proteins such as CRP, it may be that EpiScores for protein levels provide a more stable reflection of chronic inflammation. Additionally, the CRP EpiScore was found to have an average 6.4-fold stronger effect estimates in associations with brain imaging measures, versus measured CRP (10). EpiScores for CRP and IL6 inversely associated with general cognitive function in studies where the measured protein association was less strong/significant (9-11). These studies suggest that protein EpiScores might represent useful markers of brain health.

Gadd *et al.,* trained 84 protein EpiScores in the German cohort KORA which had a Pearson correlation (r) > 0.1 and P < 0.05 when compared with measured protein levels in a test cohort (12). Several of these EpiScores were found to associate with a number of disease outcomes including stroke, type 2 diabetes and lung cancer (12).

In this study, we examined if the same 84 EpiScores were associated with a general factor for cognitive function, longitudinal cognitive change, and magnetic resonance imaging (MRI) measures of global brain health and longitudinal brain changes in up to three independent cohorts (depending on data availability): Generation Scotland (GS), the Lothian Birth Cohorts of 1921 (LBC1921) and 1936 (LBC1936). We also investigated if the EpiScores associated with an incident (binary) dementia diagnosis and time-to-dementia (**Figure 1**).

**Figure 1:**
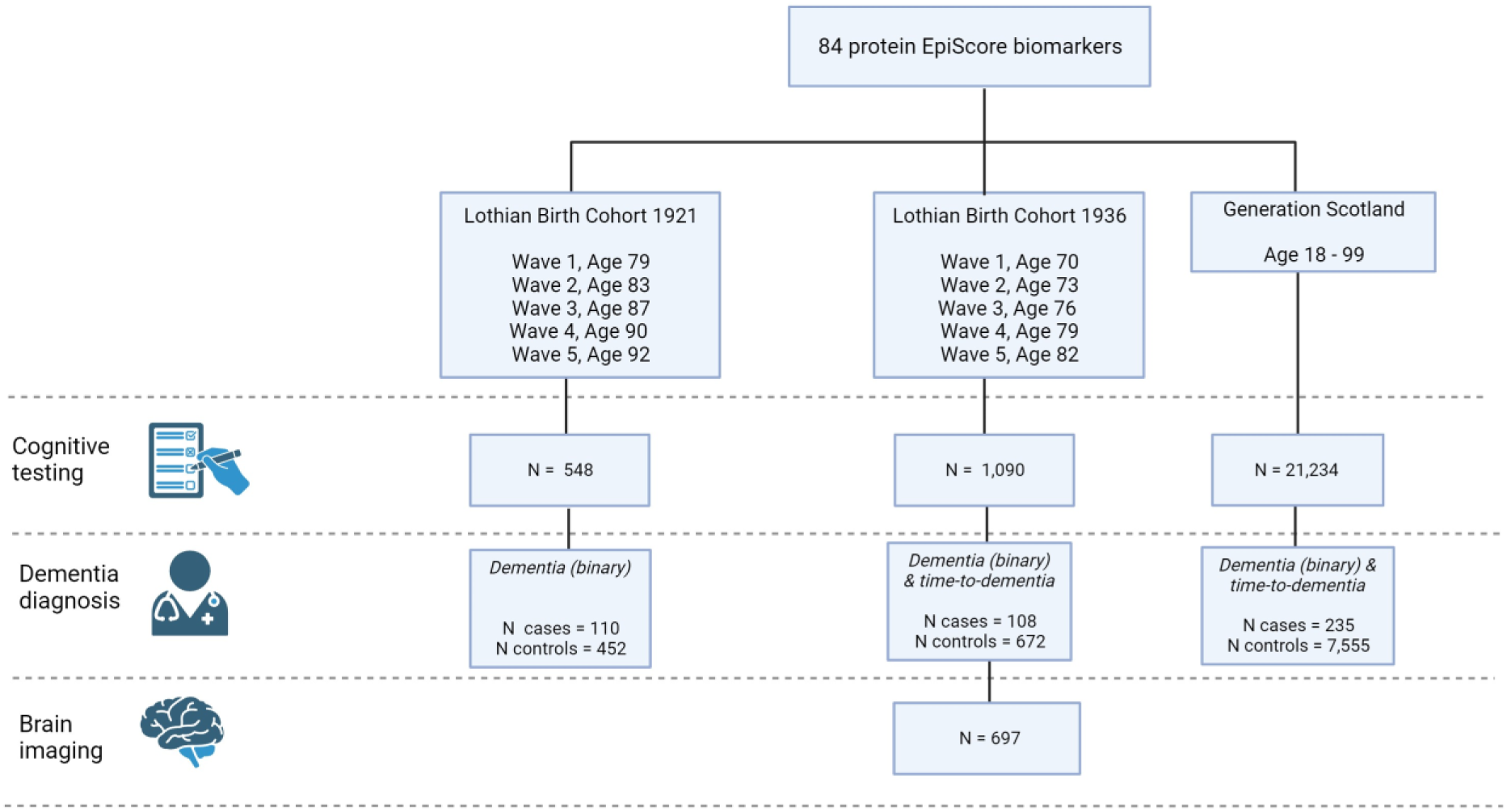
Study overview. A study summary figure highlighting the data available for cognitive testing (maximum N for one cognitive test at wave 1), dementia diagnosis (N for cases and controls with methylation data) and brain imaging (maximum N for one MRI measure at wave 2) across the LBC1921, LBC1936 and GS cohorts. Created with BioRender.com.

## Methods

### The Generation Scotland cohort

The Generation Scotland: Scottish Family Health Study (GS) has been previously described in detail by Smith *et al,* 2006 (13). In brief, GS is a cohort study of > 20,000 individuals and their families living in Scotland. GS provides a resource with genome-wide genetic, epigenetic, clinical, lifestyle and sociodemographic data. Participants in GS were aged between 17 and 99 years at the study baseline, with a mean age of 47.5 years (SD: 14.93). 58.8 % of the GS cohort is female. Recruitment took place between 2006 and 2011 (N > 20,000).

### Lothian Birth Cohorts of 1921 and 1936

The Lothian Birth Cohorts of 1921 and 1936 (LBC1921 and LBC1936) comprise older community-dwelling adults born in 1921 and 1936 (14, 15). Most of these individuals sat a test of general intelligence - the Moray House Test No.12 - at about age 11 years while at school in Scotland in 1932 and 1947, respectively. Subsequently, individuals residing in the Lothian area later in life were invited to join the LBC studies (at age ∼ 79 for LBC1921 and age ∼ 70 for LBC1936). Participants underwent a series of physical, cognitive and medical assessments at regular intervals (age ∼ 79, 83, 87, 90, 92 for LBC1921, and age ∼ 70, 73, 76, 79, and 82 for LBC1936). The participants provided blood samples from which genetic, epigenetic and biomarker data were obtained. Beginning at the second assessment (age 73), LBC1936 participants also underwent whole brain structural MRI scans. The mean age at wave 1 in the LBC1936 is 69.5 years (SD: 0.83) and 49.77% of the cohort is female. The mean age at wave 1 in the LBC1921 is 79.1 (SD: 0.58) and 58.17% of the cohort is female.

### EpiScores in the Generation Scotland and the Lothian Birth Cohorts

The training and testing of the 84 EpiScores used in this study have been described previously (12). Briefly, the 84 EpiScores are the result of penalised regression models (one model for each protein) that select CpG sites that, in weighted combination, are predictive of individual protein levels. These 84 EpiScores met a testing threshold of Pearson r > 0.1 and p < 0.05 when projected into a subset of the GS cohort (STRADL: N = 778 (16)) and compared with measured protein levels (12). EpiScores were projected into methylation data (beta values) in the LBC’s (n_LBC1921_ = 436; n_LBC1936_ = 895) and the GS cohort (n = 18,413) before being corrected for technical covariates through linear regression. Details of DNAm profiling and processing are detailed in **Appendix 1**. In GS, EpiScores were corrected for set and batch. In LBC1921 and LBC1936, EpiScores were corrected for set, array and hybridization date. Residuals from these regression models were extracted and used for all downstream analyses.

### Cognitive test data

Cognitive testing in the Generation Scotland cohort and LBC studies have been described previously (13-15, 17). Briefly, cross-sectional scores are available for four tests in GS, while longitudinal data were considered for 13 tests in LBC1936 and for four tests in LBC1921 (full details in **Appendix 1** with summary data presented in **Supplementary Tables 1-3**).

### MRI measures of brain health in LBC1936

Protocols for magnetic resonance imaging (MRI) acquisition and processing carried out in the LBC1936 cohort have been described previously (18). Four measures of global brain health were considered: total brain volume, grey matter volume, normal appearing white matter volume, and white matter hyperintensity volume. These were assessed across four waves of data collection, starting at wave 2 (age 73). Intracranial volume was included as a covariate for baseline (intercept) analyses to account for any previous volume loss. Full details are presented in **Appendix 1** with summary data in **Supplementary Table 4**.

### Dementia diagnosis information

Dementia diagnosis data was obtained in all three cohorts. Full details are provided in **Appendix 1**. Briefly, GS data were obtained via linkage to primary and secondary care records (235 incident cases, 7,555 controls – filtered so all were aged 65 or above at the time of diagnosis/censoring, **Supplementary Table 5**).

Dementia diagnosis information for LBC1921 and LBC1936 were obtained through electronic heath record (EHR) review (19). Clinician home visits were also carried out by request in LBC1921 and LBC1936 when a participant showed signs of cognitive impairment, self-reported dementia, or an LBC researcher suspected the participant may have dementia. Consensus meetings were held to discuss each participant and determine whether they had dementia, probable dementia, possible dementia or had no dementia diagnosis, as well as dementia subtype (where possible) (19). Of the participants with methylation data, there were 108 and 110 participants with a dementia diagnosis (692 and 452 controls) in LBC1936 and LBC1921, respectively (**Supplementary Table 5**). Date of diagnosis/time-to-event information was only available in LBC1936.

### Statistical analysis

All statistical analysis were performed in R version 4.0.3 (2020-10-10) (20).

#### Descriptive Statistics

Sample sizes for cognitive, brain MRI measures and dementia shown in **Figure 1** highlight the maximal data available. Sample sizes vary across tests and decrease over follow-up in both LBC cohorts. Therefore, data available for each test/measure at each wave can be found in **Supplementary Tables 1-5**.

### Predictors of cognitive function, cognitive change and MRI brain health measures

All analyses in this study included basic- and fully-adjusted models. Outcomes of interest were latent intercept and slope variables for brain and cognitive outcomes (see **Appendix 1** for details and **Supplementary Tables 6 - 9**). Regression analyses were performed within the structural equation framework. Continuous covariates were scaled to aid in model convergence and to obtain standardized regression coefficients.

*Basic model: Outcome of interest ∼ EpiScore + Age at baseline + Sex*

*Full model: Outcome of interest ∼ EpiScore + Age at baseline + Sex + Scottish Index of Multiple Deprivation (SIMD) + Epigenetic smoking score (EpiSmoker) + Body Mass Index (BMI) + Alcohol units per week*

Information regarding alcohol intake (weekly units) was obtained via a self-reported questionnaire. The Scottish Index of Multiple Deprivation (SIMD, 2006) in LBC1936 and GS, and social grades determined by highest reached occupation in LBC1921 (21, 22). The SIMD ranged from 1 (most deprived) – 6505 (least deprived). Body Mass Index (BMI in kg/m^2^) was obtained via an in-clinic physical assessment.

Epigenetic smoking scores were calculated for each participant from their DNAm profiles using the R package *EpiSmokEr (23)*.

Descriptive statistics for all covariates in GS, LBC1936 and LBC1921 can be found in **Supplementary Tables 10 – 12**.

### Dementia analysis

Associations between the EpiScores and incident dementia (binary outcome) were tested in all three cohorts using logistic regression models with the “glm” function (with family set to binomial) from the R *stats* package (version: 4.0.3) (20). Time-to-dementia analyses were also run in LBC1936 and GS using Cox proportional hazards (CoxPH) models through the R *survival* package (version: 3.3.1) (24). Sensitivity analyses to account for related individuals (GS) and death as a competing risk (GS and LBC1936) were also considered (details in **Appendix 1**).

In GS, baseline appointments were from 2006 to 2011 and the dementia censor date was set to April 2022 resulting in a maximum of ∼ 11 – 16 years lag time between sample collection and dementia. In LBC1936, sample collection was carried out at baseline appointment where participants were ∼ age 70 and maximum age at the last dementia ascertainment is 86.44 years resulting in a maximum lag time of 16.44 years between sample collection and dementia. In LBC1921, sample collection was carried out at baseline appointment where participants were ∼ age 79.1 years. The consensus meeting was in 2016 meaning the maximum age at dementia diagnosis could be 95, therefore the maximum lag time between sample collection and dementia is ∼ 16 years.

### Meta-analyses

Meta-analyses was performed to obtain effect sizes weighted by sample size using results from the general cognitive function, dementia diagnosis (binary) and time-to-dementia models using the R package *metafor* (version: 4.2-0) (25).

## Results

### EpiScore associations with General cognitive function

A latent factor of general cognitive function (intercept) generated in three separate cohorts was regressed on 84 EpiScores in separate linear models. Benjamini and Hochberg false discovery rate (P_FDR_ < 0.05) correction was applied to results to account for multiple testing. In the basic models (adjusted for age and sex), 20 (GS), 13 (LBC1921) and 31 (LBC1936) EpiScores were significantly associated with general cognitive function (Absolute standard effect size range: 0.09 – 0.41, P_FDR_ < 0.05, **Supplementary Table 13**). Fully-adjusted models were also examined in which no significant associations were found in LBC1936, 5 associations were found in LBC1921, and 40 associations in GS (Absolute standard effect size range: 0.02 – 0.53, P_FDR_ < 0.05, **Supplementary Table 13**). A meta-analysis of effect sizes for general cognitive function in all three cohorts was performed for basic- and fully-adjusted model results (**Supplementary table 14**). In the meta-analysis of the basic results for general cognitive function, 36 EpiScores were found to be significantly associated (Absolute standard effect size range: 0.06 – 0.22, P_FDR_ < 0.05). 18 EpiScore associations from the fully-adjusted models were significant (Absolute standard effect sizes range: 0.03 – 0.14, P_FDR_ < 0.05, **Figure 2**).

**Figure 2:**
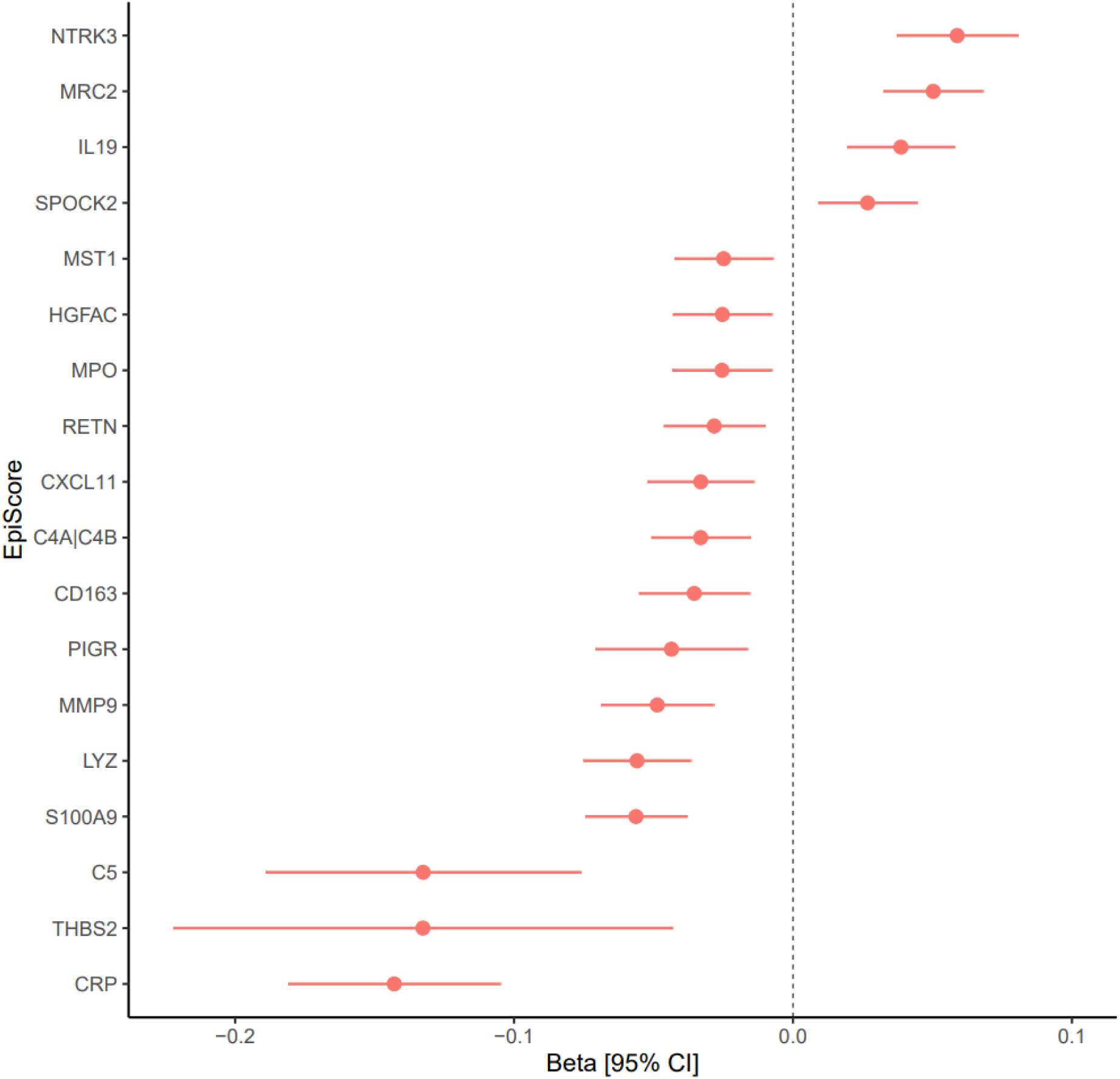
Meta-analysis of EpiScore associations with general cognitive function in three cohorts. The plot shows the meta-analysed regression coefficients for each EpiScore from the fully adjusted models, found to be significantly associated with general cognitive function after FDR correction. Error bars indicate 95% confidence intervals [95% CI].

Next, in LBC1921 and LBC1936, a general latent factor of cognitive change (slope) was regressed on the 84 EpiScores in separate linear models. No EpiScores were significantly associated with a general factor of cognitive change in either LBC1921 or LBC1936 in models with basic adjustments after FDR correction. However, three EpiScores were nominally associated (Absolute standard effect size range: 0.19 – 0.2, P < 0.05) with slope in the LBC1921. Fully-adjusted models were also examined in which no FDR significant associations where found in either cohort (**Supplementary Table 15)**. One and three EpiScores were nominally associated with slope in LBC1936 and LBC1921, respectively (Absolute standard effect size range: 0.09 – 0.25, P < 0.05) in the fully adjusted models. The number of associations with cognitive function and change summarised in **Supplementary Table 16**.

### EpiScore associations with MRI measures of global brain health

The 84 EpiScores were then studied in relation to four MRI markers of brain health (total brain volume, grey matter volume, normal appearing white matter volume, and white matter hyperintensity volume) and their changes over time in LBC1936. In basic models adjusted for age and sex, 21 EpiScores were significantly associated with total brain volume, 28 with grey matter volume, 16 with normal appearing white matter volume and 3 with white matter hyperintensity volume (Absolute standard effect size range: 0.04 – 0.21, P_FDR_ < 0.05). Eleven EpiScores were found to associate with three or more MRI measures of brain health in the basic models (P_FDR_ < 0.05, **Figure 3**). Fully adjusted models were examined to determine if associations were attenuated when covariates relevant to brain health were included in the model (**Supplementary Table 17**). One EpiScore, CRP, was found to be associated with grey matter volume at baseline (Standard effect size: - 0.09, P_FDR_ < 0.05).

**Figure 3:**
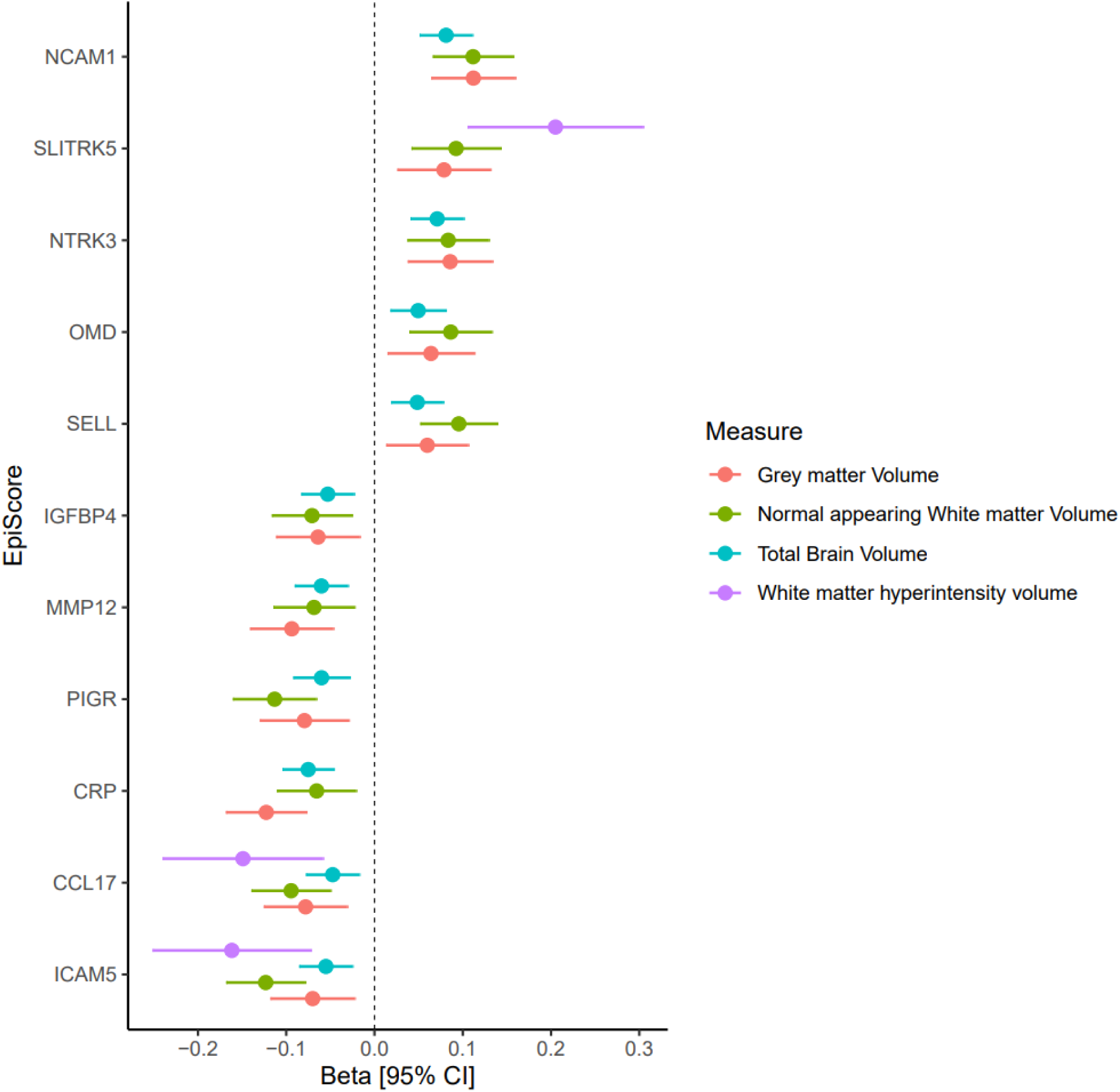
EpiScore associations with cross-sectional MRI measures of brain health in the LBC1936 cohort. Plot shows the standardised regression coefficients for each EpiScore found to be significantly associated with three or more MRI measures of brain health in basic models in the LBC1936 cohort (FDR < 0.05). Error bars indicate 95% confidence intervals [95% CI]. Direction of effect sizes and 95% CI have been recoded for white matter hyperintensity volume.

EpiScore associations with the slope (change over ∼ 9.5 years) for each MRI measure were tested; no FDR significant results were observed. However, nominally significant associations were observed for all four measures in models with basic adjustments (Absolute standard effect size range: 0.12 – 0.3, P < 0.05, **Supplementary Table 18**). A summary table for the number of associations observed with cross-sectional and longitudinal MRI measures for basic and fully-adjusted models can be found in **Supplementary Table 19**.

### EpiScore associations with incident dementia

EpiScores associations with a binary dementia diagnosis and time-to-dementia were examined. As age at dementia diagnosis was not available in the LBC1921, this cohort was only included in logistic regression models testing the binary outcome for dementia. In the logistic regression models with basic adjustments, three significant associations: SEMA3E (OR: 1.54), ICAM5 (OR: 0.66), and PIGR (OR: 0.66) were observed in LBC1921 (P_FDR_ < 0.05). Of these associations, the ICAM5 EpiScore (OR: 1.2) was nominally significant in GS (P < 0.05). The remaining two associations were not nominally significant in GS or LBC1936 (**Figure 4 - Panel A, Supplementary table 20**).

**Figure 4:**
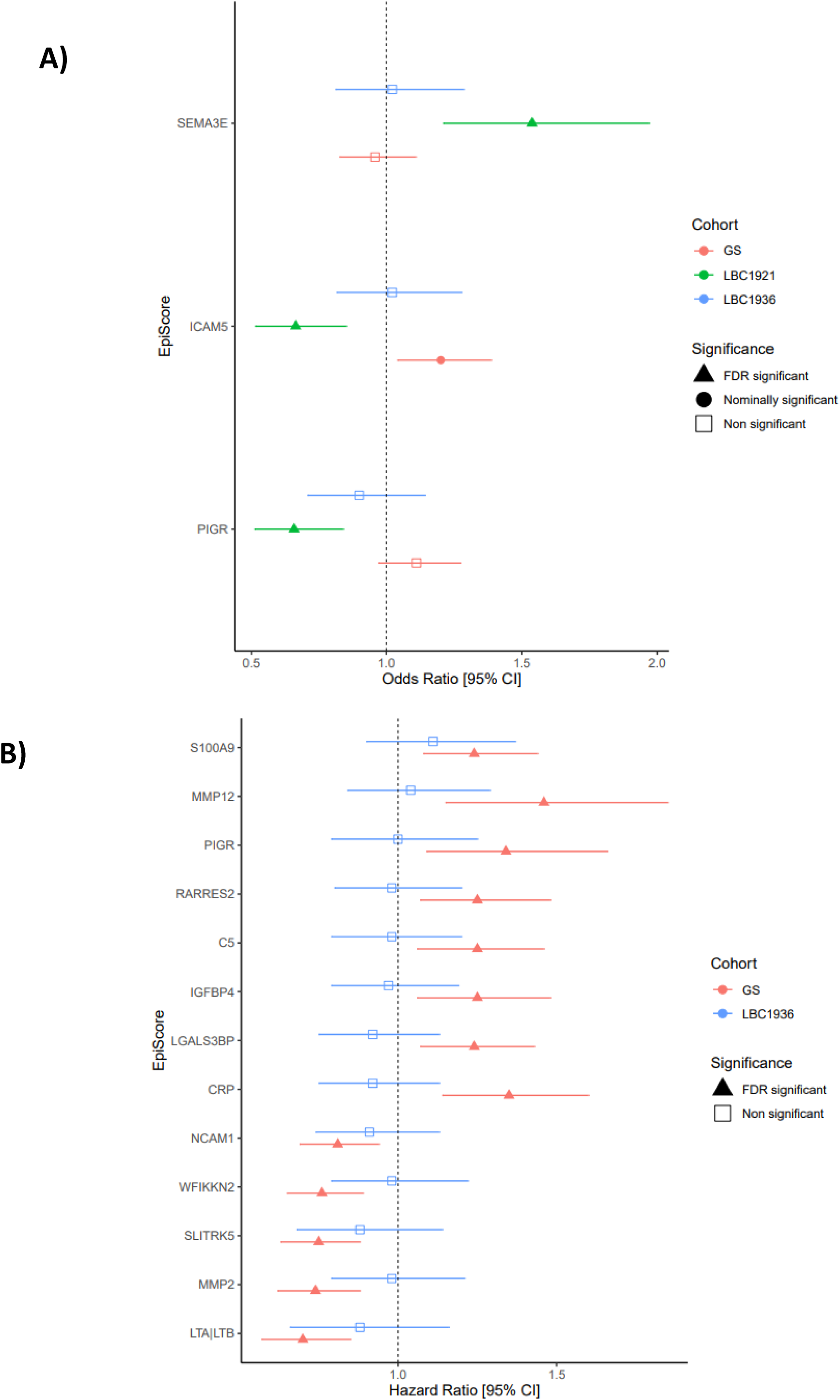
EpiScore associations with incident dementia (binary) and time-to-dementia. Panel A: FDR significant Odds Ratios for EpiScores with dementia status (binary) in LBC1921. The Odds Ratios for GS and LBC1936 for the same EpiScores have been included for comparison despite being only nominally significant or non-significant. Panel B: FDR significant Hazard Ratio for EpiScores with incident dementia for the mixed effects Cox models in GS. The Hazard Ratios for LBC1936 from the CoxPH model for the same EpiScores have been included for comparison despite being non-significant (Panel B). All error bars represent 95% confidence intervals [95% CI].

CoxPH models were used to test the association between EpiScores and time-to-dementia in GS and LBC1936 (**Supplementary table 21**). Additionally, mixed effects Cox models were run in GS to account for relatedness (**Supplementary table 22**). In the basic mixed effects models for GS, 13 significant (P_FDR_ < 0.05) associations were observed; these were not found to be significant in the LBC1936 cohort (**Figure 4 - Panel B**). Only the MMP2 EpiScore was significant (HR: 0.71, P_FDR_ < 0.05) after full adjustments were made to the mixed effects models in GS. No significant findings were observed in the competing risk models for either cohort (**Supplementary Table 23**). However, there was good agreement between the hazard ratios from the cause-specific and competing risk models (**Supplementary Figure 4).** Additionally, separate meta-analyses of the results obtained from the logistic regression models and the time-to-event analyses were carried out (**Supplementary Tables 24-25**). No EpiScores were found to be significant in either analysis after FDR correction.

## Discussion

In this study, we identified multiple associations between protein EpiScores and measures of cognitive function, MRI proxies of brain health and dementia in three independent cohorts.

### EpiScore associations with general cognitive function and global brain volume

Eighteen EpiScores were significantly associated with general cognitive function in the meta-analysis of the fully-adjusted results. Three of these EpiScores (for CRP, PIGR, and NTRK3) were also associated with total brain volume, grey matter volume, and normal appearing white matter volume at baseline in models with basic adjustments. Furthermore, the EpiScore for PIGR also was associated with incident dementia (as a binary outcome) in the LBC1921 basic-adjusted model (OR: 0.66, P_FDR_ < 0.05) and time-to-dementia in the GS mixed effects Cox models with basic adjustments (HR: 1.34, P_FDR_ < 0.05). The EpiScore for CRP was also found to associate with time-to-dementia in the GS mixed effects model with basic adjustments (HR: 1.35, P_FDR_ < 0.05).

CRP is an acute-phase inflammatory protein, mainly transcribed in response to high levels of inflammatory proteins (26-28). In previous studies performed with the LBC1936 and GS, an EpiScore for CRP was found to be negatively associated with cognitive function (9, 10). Differences in methodology between this study and previous studies in LBC1936 and GS exist. However, this comparison, particularly in GS where the sample size is over an order of magnitude greater than previous CRP EpiScore – cognition studies, provides excellent replication for the association. To our knowledge, no association between PIGR or NTRK3 with general cognitive function/global volumetric MRI measures of brain health have been described previously. PIGR is expressed in the endothelial cells of the blood-brain barrier and binds to the bacteria *Streptococcus pneumonia (Pneumococci)* – a leading causes of bacterial meningitis (29). According to the World Health Organisation, 1 in 5 individuals who previously had meningitis suffer from long-term complications including cognitive impairments (30). NTRK3 binds Neurotrophin-3, an important neuro-growth factor. A reduction of transcript levels of NTRK3, also known as tyrosine kinase receptor C (trkC), has been observed in patients with schizophrenia (31, 32). Previous studies have also highlighted a potential association between NTRK3 and hippocampal function in both mice and humans (33-35).

### S100A9 EpiScore associates with time-to-dementia in GS

Thirteen EpiScores were significantly associated with incident time-to-dementia in the GS Cox mixed effects model (basic adjustments). Of these EpiScores, four (PIGR, S100A9, C5, CRP) were found to overlap with associated EpiScores in the meta-analyses of the general cognitive function results (fully adjusted models). Seven EpiScores (NCAM1, SLITRK5, IGFBP4, MMP12, PIGR, CRP, ICAM5) overlapped with associations observed in three or more of the MRI global volumetric measures at baseline in LBC1936 (basic adjustments). The S100A9 EpiScore is of particular interest as it has been previously identified as a potential biomarker of Alzheimer’s disease (36). A significant inverse association was also observed between the S100A9 EpiScore and general cognitive function in the meta-analysis, in the fully-adjusted model. S100A9 is known to co-localise with amyloid beta and is thought to contribute to plaque formation (36, 37). A reduction of S100A9 in an Alzheimer’s disease mouse model resulted in less amyloid beta plaques and less cognitive impairment (38). In cerebrospinal fluid of patients with Alzheimer’s disease, significantly lower levels of S100A9 protein has been observed compared with controls (36). The lack of replication observed in the LBC1936 could be due to using all-cause dementia as a phenotype and biomarkers may be specific to certain subtypes of dementia. Future work could investigate the subtypes of dementia to determine if different EpiScores associate with a specific subtype.

### Strengths and limitations

Strengths of this study include the large sample sizes and multi-cohort analyses. Further, inclusion of the longitudinal Lothian Birth Cohorts facilitated the study of EpiScores as biomarkers of cognitive change over time. Different sample sizes and lifestyle factors as well as age profiles may explain why some EpiScore associations did not replicate across all cohorts. However, the scores that did replicate across all cohorts are potential biomarkers of cognitive function across the mid-to-late life.

A limitation of this study is that the population base is of European ancestries and living in Scotland and so may not generalise to other populations. Further work is needed to investigate if the findings are generalisable across the life course. This is important to understand because early life immune dysregulation contributes to some neurodevelopmental disorders. For example, a recent study found that a DNAm-based proxy of CRP correlates with inflammation burden and MRI markers of encephalopathy of prematurity after preterm birth (39). Another limitation of this study is the lack of replication for MRI findings and the consideration of EpiScores from a single time-point. The absence of the measured proteins in these cohorts is also a limitation as we were unable to compared EpiScore performance against measured protein.

### Conclusion

In conclusion, 84 protein EpiScores were tested against measures of general cognitive function, brain health and incident dementia across three human cohorts. Several EpiScores analysed in this study may augment typical risk factors of brain health, however further replication studies are required.

## Ethics declarations

All components of GS received ethical approval from the NHS Tayside Committee on Medical Research Ethics (REC Reference Number: 05/S1401/89). GS has also been granted Research Tissue Bank status by the East of Scotland Research Ethics Service (REC Reference Number: 20-ES-0021), providing generic ethical approval for a wide range of uses within medical research.

Ethics permission for the Lothian Birth Cohort 1936 (LBC1936) was obtained from the Multi-Centre Research Ethics Committee for Scotland (Wave 1: MREC/01/0/56), the Lothian Research Ethics Committee (Wave 1: LREC/2003/2/29), and the Scotland A Research Ethics Committee (Waves 2, 3, 4 & 5: 07/MRE00/58).

Ethics permission for the Lothian Birth Cohort 1921 (LBC1921) was obtained from the Lothian Research Ethics Committee (Wave 1: LREC/1998/4/183; Wave 2: LREC/2003/7/23; Wave 3: LREC1702/98/4/183) and the Scotland A Research Ethics Committee (Wave 4: 10/S1103/6; Wave 5: 10/MRE00/87).

## Data availability

The source datasets from the cohorts that were analysed during the current study are not publicly available due to them containing information that could compromise participant consent and confidentiality. Data can be obtained from the data owners. Instructions for accessing Generation Scotland data can be found here: https://www.ed.ac.uk/generation-scotland/for-researchers/access; the ‘GS Access Request Form’ can be downloaded from this site. Completed request forms must be sent to to be approved by the Generation Scotland Access Committee. According to the terms of consent for GS participants, access to data must be reviewed by the GS Access Committee. Instructions for accessing Lothian Birth Cohort data, alongside a Data Request Form template, Data Summary Tables and Data Dictionaries can be found here: https://www.ed.ac.uk/lothian-birth-cohorts/data-access-collaboration.

## Code availability

All R code used in analyses is provided at: https://github.com/hmsmith22/EpiScore_biomarkers_of_brain_health

## Author contributions

H.M.S., R.E.M., J.E.M., and S.R.C. were responsible for the conception and design of the study. H.M.S. carried out the data analyses. H.M.S., drafted the article. A.D.C, K.M.G., R.F.H., and D.L.Mc., contributed to methodology. A.C., M.V.H., S.M.M., M.E.B., J.M.W., J.C., A.T., D.P., I.J.D. contributed to the data collection and preparation. R.E.M., S.R.C., and J.E.M. supervised the project. All authors read and approved the manuscript. Competing interests R.E.M is an advisor to the Epigenetic Clock Development Foundation, and Optima partners. R.F.H. has received consultant fees from Illumina and Optima partners. D.A.G. has received consultant fees from, and is currently employed in part-time capacity, by Optima Partners. D.L.McC. and is currently employed in part-time capacity, by Optima Partners. All other authors declare no competing interest.

## Materials and correspondence

All correspondence and material requests should be sent to Riccardo Marioni at.

