## Supplementary Figures for "Epigenetic scores of blood-based proteins as biomarkers of general cognitive function and brain health"

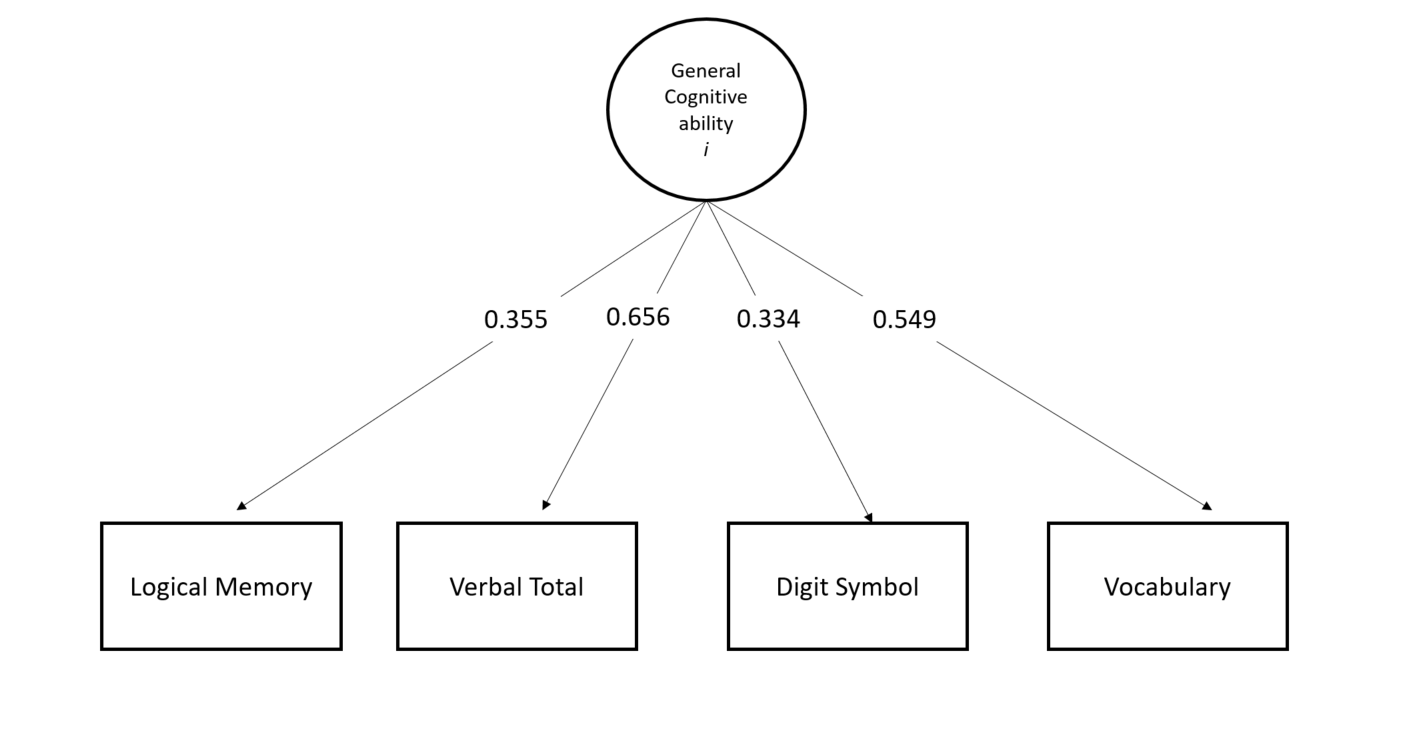

**Supplementary figure 1: General Cognitive ability in GenScot.** Path diagram describing the measurement model of general cognitive ability in GenScot. Model fit measures can be found in Supplementary table 6 and loadings in Supplementary table 7.

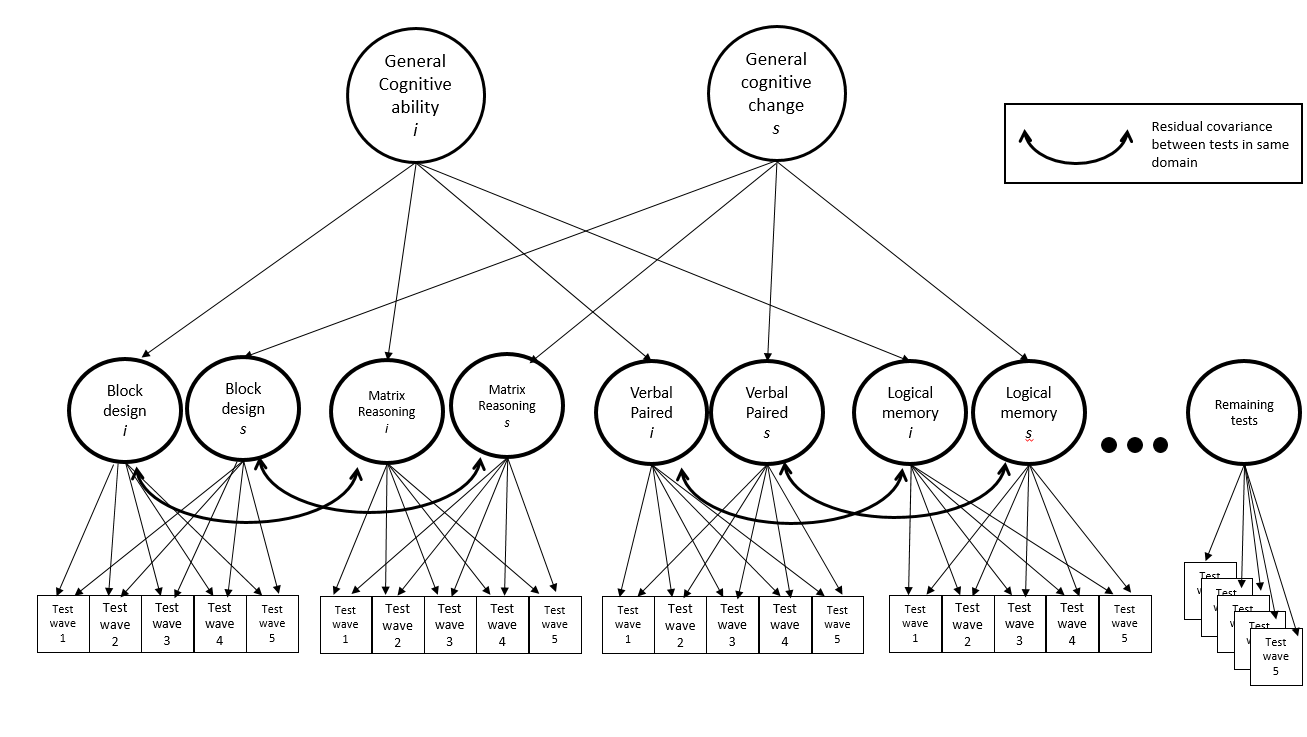
**Supplementary figure 2: General Cognitive ability in LBC1936.** Path diagram describing the measurement model of general cognitive ability and change in LBC1936. Model fit measures can be found in Supplementary table 6 and loadings in Supplementary table 7.

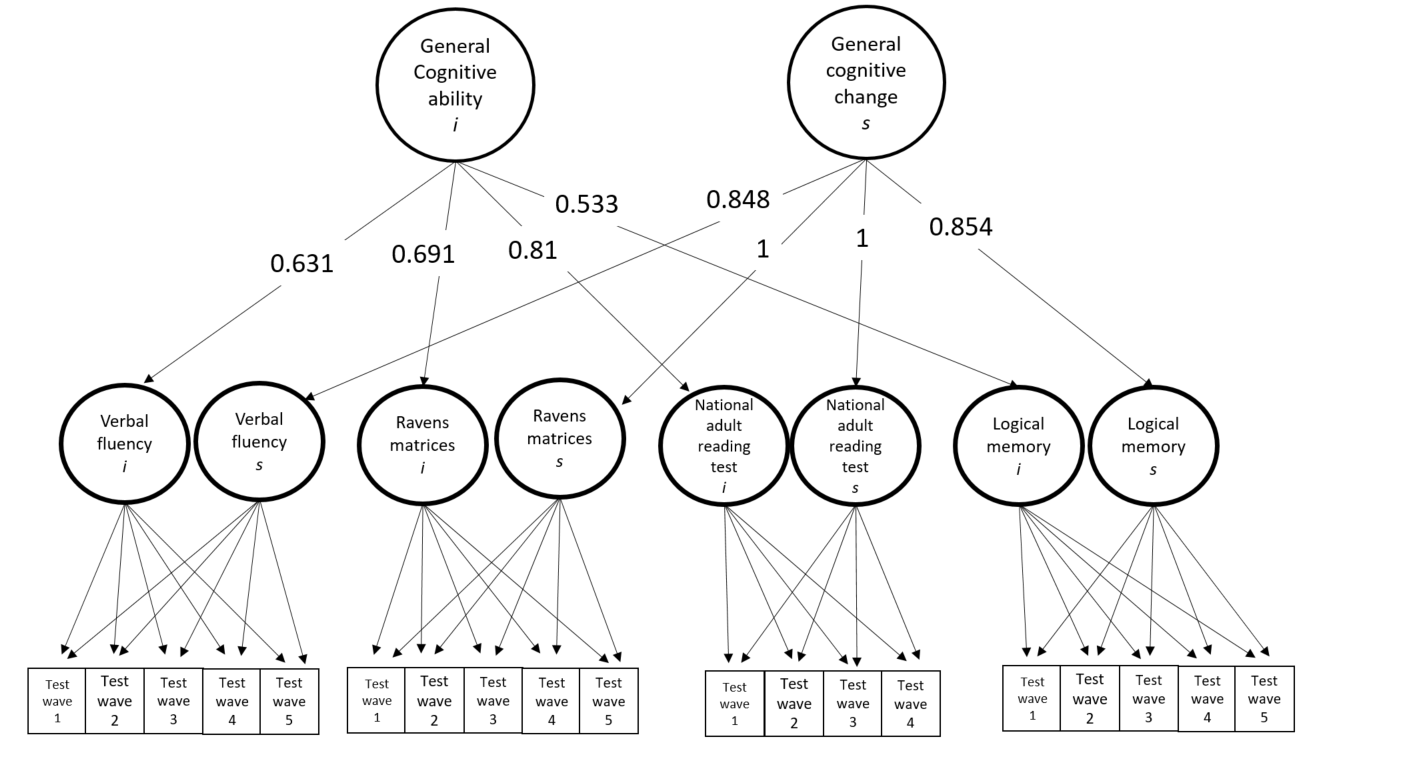

**Supplementary figure 3: General Cognitive ability in LBC1921.** Path diagram describing the measurement model of general cognitive ability and change in LBC1921. Model fit measures can be found in Supplementary table 6 and loadings in Supplementary table 7.

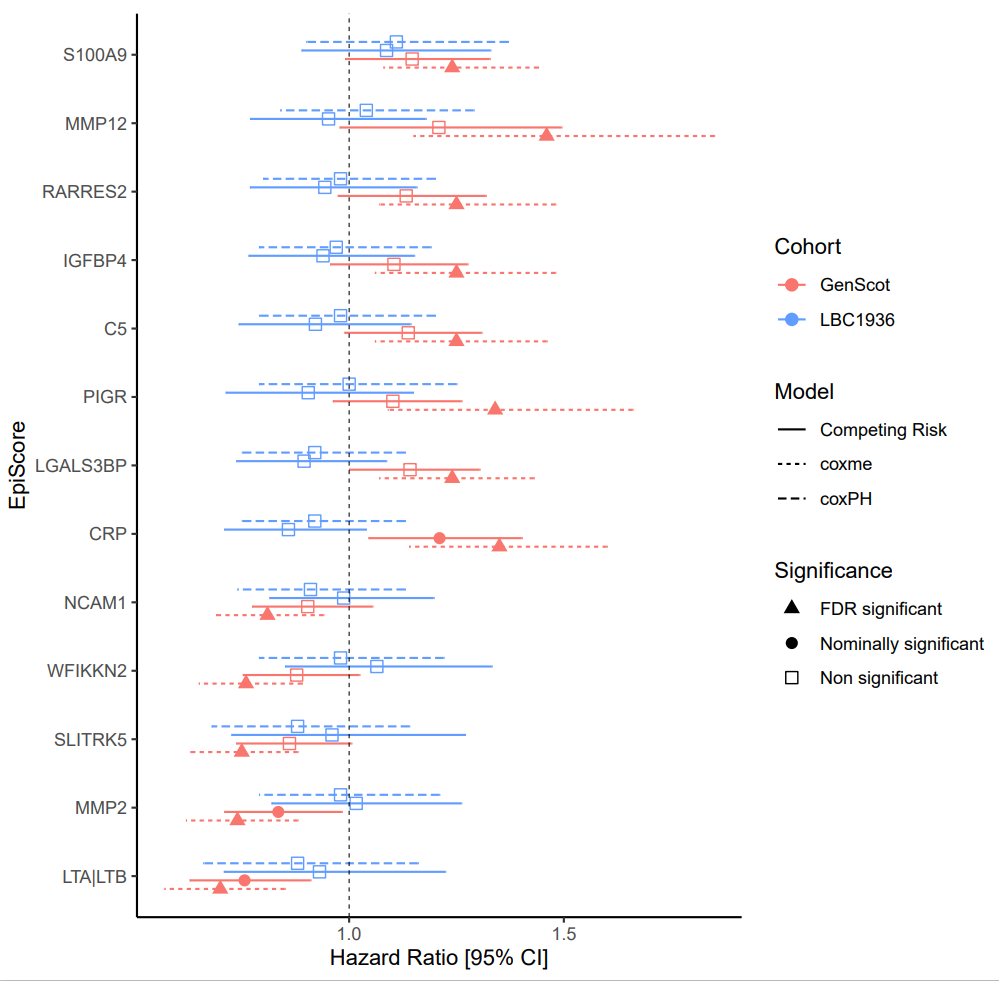

**Supplementary figure 4: Time-to-dementia in GS and LBC1936.** FDR significant Hazard ratio for EpiScores with incident dementia for the mixed effects Cox models in GS. The Hazard ratios for LBC1936 from the coxPH model for the same EpiScores have been included for comparison despite being non-significant. The Hazard ratios for the competing risk models for GS and LBC1936 are also shown for comparison. All error bars represent 95% confidence intervals [95% CI].
