## Appendix 1 for "Epigenetic scores of blood-based proteins as biomarkers of general cognitive function and brain health"

**DNAm data in Generation Scotland and the Lothian Birth Cohorts**

In GS, DNA was extracted from blood samples of > 18,000 individuals and DNAm analysed using the Illumina EPIC array (850K). The analyses were run in multiple sets: set 1 (N = 5,087), set 2 (N = 4,450), set 3 (N = 8,876). A subset of individuals in set 1 are a mixture of biological and non-biological relatives of each other. Set 2 participants are not related to the individuals in set 1, nor are they related to each other. Set 3 participants are related to each other and/or are related to participants in sets 1 and 2. Quality control (QC) of DNAm data was performed to remove outliers and identify any errors during data acquisition. A detailed description of QC has been described previously (1-3). Briefly, individuals who responded “yes” to all self-reported disease questions in health questionnaire were excluded from analysis (N = 3). Outliers and sex mismatches were removed from analysis. Non-blood samples i.e. salvia sample were excluded. Poor quality samples including samples with missing CpG values and/or poorly-detected CpGs were removed. Non-autosomal and non-CpG sites were excluded from analysis.

In LBC1921 and LBC1936, DNA was extracted from blood samples and DNAm was analysed using the Illumina methylation array (450K). QC of DNAm data has been described elsewhere (4-6). In brief, original intensity data were background-corrected and normalised to controls. Low quality samples were manually removed due to technical errors including insufficient hybridization or nucleotide extension and problems with bisulfide conversion or signal staining. Probes with low detection and call rates were removed (detection rate < 95% and probes detected < 450,000 at p < 0.01). Samples with incorrect DNAm predicted sex and mismatches between genotype and SNP probes were removed (4).

**Cognitive measures in the Generation Scotland cohort**

Cross-sectional test scores are available for four tests: Digit Symbol Substitution Test, Verbal Fluency Fest, Vocabulary and Logical Memory (7). The Logical Memory variable was created by combining two test scores for immediate and delayed recall. Any test scores beyond 3.5 standard deviations from the mean were considered outliers and recoded as missing. Data availability and descriptive statistics for each cognitive test can be found in **Supplementary Table 1**.

**Cognitive measures in the Lothian Birth Cohort 1936**

Cognitive testing in the Lothian Birth Cohorts has been described previously (8-11). In the LBC1936 cohort, scores for thirteen cognitive tests were available across 5 waves (age 70 (baseline), 73, 76, 79, 82).The Block Design, Matrix Reasoning (WAIS-III^UK^) and Spatial Span (WMS-III^UK^) tests were used to measure visuospatial ability. The National Adult Reading Test, Wechsler Adult Reading Test and Verbal Fluency Test (using letters C, F and L) were all used to examine verbal ability. The Verbal Paired Associates, Logical Memory – a combination of immediate and delayed memory (WMS-III^UK^) and Digit Span Backwards (WAIS-III^UK^) tests were used to assess memory. The Digit Symbol Substitution Test, Symbol Search (WAIS-III^UK^), Choice Reaction Time and Inspection Time were used to evaluate processing speed. Data availability and descriptive statistics for each cognitive test at respective waves can be found in **Supplementary Table 2**.

**Cognitive measures in the Lothian Birth Cohort 1921**

In LBC1921, results for the Verbal Fluency Test, Logical Memory (Wechsler Memory Scale—Revised) and the Ravens Progressive Matrices tests were available across five waves (~ age 79 (baseline), 83, 87, 90, 92). Results for the National Adult Reading Test (NART) were available across four waves (~ age 79 (baseline), 83, 86, 90). The NART and Verbal Fluency Test assessed verbal ability, whilst Ravens Progressive Matrices assessed nonverbal reasoning ability. Logical Memory tests assessed both immediate and delayed recall. Data availability and descriptive statistics for each cognitive test can be found in **Supplementary Table 3**.

**MRI measures of brain health in LBC1936**

Protocols for magnetic resonance imaging (MRI) acquisition and processing carried out in the LBC1936 cohort have been described previously (12). Briefly, at the Brain Research Imaging Centre at the University of Edinburgh, a 1.5T GE Signa Horizon HDx clinical scanner was used to collect T_1_-weighted (voxel size: 1 x 1 x 1.3 mm) T_2_-weighted (voxel size: 1 x 1 x 2 mm), T_2_*-weighted (voxel size: 1 x 1 x 2 mm), and fluid-attenuated inversion recovery-weighted images (FLAIR) (voxel size: 1 x 1 x 4 mm) (12). The following structural MRI measures were used as a proxy for global brain health including total brain volume, grey matter volume, normal appearing white matter volume and white matter hyperintensity volume. MRI measures were available across four waves of the LBC1936 (waves 2 – 5) which extends across a ~ 9.5 year period. Data availability and descriptive statistics for MRI measures can be found in **Supplementary Table 4**.

**Dementia diagnosis information**

*Generation Scotland*

Dementia diagnosis data in the GS cohort was available through electronic health records data linkage. Of the participants whose data was included in this part of the study, despite all volunteers providing consent for access to their GP data, not all GP surgeries agreed to release their data. In total, GP records were available for 7,580 individuals and hospital records were available for 21,725 individuals. A censoring date was set at April 2022 – the latest date for which hospital data were available. The first report of dementia in a participant’s records was used as the date of dementia diagnosis. If an individual did not have a report of a dementia diagnosis prior to or in October 2020 in their hospital records (or GP records for the sub-group from whom data were available) then they were allocated to the control group (i.e. non-dementia group). Due to age at baseline ranging from 18 upwards in GS, participants were filtered to age at event (dementia or censor) ≥ 65 years (to only consider late onset dementia). Of the participants with methylation data, 235 individuals had all-cause dementia diagnosis whilst 7,555 individuals did not have a dementia diagnosis at censor (**Supplementary Table 5**). Time to event was calculated by subtracting age at baseline study appointment from age at diagnosis/censor.

*LBC1936*

Dementia diagnosis information for the LBC1936 was obtained through electronic heath record (EHR) review (13). Clinician home visits were also carried out by request when a participant show signs of mild cognitive impairment, self-reported dementia or an LBC researcher suspected the participant may have dementia. After EHR review and home visits, a consensus meeting was held to discuss each case and determine whether a participant had dementia, probable dementia, possible dementia or had no dementia diagnosis (13). The LBC1936 participants were filtered to those present from wave 2 onwards, due to health record linkage consent starting at this wave. The censoring date for each participant was set as the date their medical records were last checked. Of the participants with methylation data, 13 participants with mild cognitive impairment, 7 participants with possible dementia, and 1 participant without linkage consent were removed from the dataset. 108 participants with methylation data had a dementia diagnosis and 692 participants did not have a dementia diagnosis at the censoring time point (**Supplementary Table 5**). Time to event was calculated using the same method as in GS.

*LBC1921*

In LBC1921, evidence of dementia diagnosis was obtained via death certificate, electronic general medical records, and electronic psychiatric records. A consensus meeting was held on the 15^th^ of December 2016 to consider the evidence for dementia diagnosis and cases were recorded as probable dementia, possible dementia or no dementia (14). 7 participants with possible dementia were removed from the dataset. 110 participants had a dementia diagnosis and 452 participants formed the control group (**Supplementary Table 5**). Age at dementia diagnosis information was not available for LBC1921, therefore it was not possible to calculate time-to-event for this cohort.

**General cognitive function**

Latent measures of general cognitive function were generated modelled using confirmatory factor analysis in a structural equation modelling (SEM) framework for each cohort, using the R package *Lavaan* (version 0.6-12) (15). Model fit measures are reported, including confirmatory factor index (CFI), Tucker-Lewis index (TLI), root mean squared error approximation (RMSEA) and the standardised root mean squared residual (SRMR) (**Supplementary Table 6**). For each cohort, the marker method was used to scale according to the first variable to aid model convergence, and all models employed full information maximum likelihood to include all data available. Negative residual variances were fixed to zero. In GS, only cross-sectional cognitive data were available, and four cognitive tests (Logical Memory, Verbal Fluency Test, Digit Symbol Substitution Test and Vocabulary) indicated a latent factor for general cognitive function (**Supplementary Figure 1,** **Supplementary Table 7**). In LBC1936 and LBC1921, levels and changes in cognitive functioning were modelled with a latent growth curve model (LGCM) using a Factor of Curves specification (16). In both cases, the intercepts and slopes of each cognitive test (thirteen cognitive tests for LBC1936, and four for LBC1921) were used to indicate a latent intercept and slopes (level and change) of general cognitive function (**Supplementary figures 2-3**, **Supplementary Table 7**). In LBC1936, a first-order hierarchical cognitive model was specified. Residual covariance between tests in the same cognitive domain were specified with reference to prior work to establish the correlational structure of cognitive domains (speed, memory, verbal ability and visuospatial (17)). In LBC1921, data was available across five waves for three out of the four cognitive tests (Logical Memory, Raven’s Progressive Matrices, and Verbal Fluency) and for four waves for the National Adult Reading Test. In both LBC cohorts, the growth curve slopes were weighted by mean lag time between each wave and baseline.

**Cross-sectional and longitudinal measures of brain health**

Growth curve models in a SEM framework were used to generate latent variables representing a baseline (intercept) and change (slope) for each MRI measure of brain health. The measurements for each brain measure were loaded onto a latent variable for intercept with loadings fixed at 1 for each wave. The measurements for each brain trait were also loaded onto a latent variable for slope with each loading fixed at the mean lag time between the corresponding wave and baseline, as above. Model fit measures for the measurement model for each MRI variable can be found in **Supplementary Table 8** and loadings can be found in **Supplementary Table 9**.

**Dementia analysis**

The usefulness of EpiScores as a biomarker of incident dementia (binary outcome) was tested in all three cohorts using logistic regression models with the “glm” function (with family set to binomial) from the *stats* package (version: 4.0.3) in R (18). We examined the EpiScores as potential biomarkers of time-to-dementia in LBC1936 and GS using Cox proportional hazards (CoxPH) models through the *survival* package (version: 3.3.1) in R (19). Additionally, in the GS cohort, EpiScore associations with time-to-dementia were tested using the *coxme* R package (version: 2.2.16) to enable adjustment for a kinship matrix (20). This method is only relevant to the GS cohort due to the presence of related individuals in the study.

Furthermore, in both GS and LBC1936 cohorts, a subset of participants died within the follow-up period. There is no way of knowing if these individuals would have gone on to be diagnosed with dementia had they not died. Statistically, this presents a difficulty as these individuals are classed as healthy/censored controls in the CoxPH/coxme models. To address this, we conducted a supplementary analysis in which we considered a sub-distribution hazard model using the *cmprsk* package (version: 2.2.11) in R (21). This tests the association between an EpiScore and the instantaneous rate of hazard for non-dementia diagnosed participants and those who have died without a dementia diagnosis.

18.	Team RC. R: A language and environment for statistical computing. MSOR connections. 2014;1.

19.	Therneau TM. A package for Survival Analysis in R. 2023.

20.	Therneau TM. coxme: Mixed Effects Cox Models. 2020.

21.	Gray B. cmprsk: Subdistribution Analysis of Competing Risks. 2022.
